# Glycosylation state of vWF in circulating extracellular vesicles serves as a novel biomarker for predicting depression

**DOI:** 10.1101/2024.03.24.24304794

**Authors:** Norihiro Yamada, Kana Tominaga, Naoomi Tominaga, Ayumi Kobayashi, Chihiro Niino, Yuta Miyagi, Hirotaka Yamagata, Shin Nakagawa

**Affiliations:** Division of Neuropsychiatry, Department of Neuroscience, Yamaguchi University Graduate School of Medicine, 1-1-1 Minami-kogushi, Ube, Yamaguchi, 755-8505, Japan; Department of Laboratory Medicine, Faculty of Health Sciences, Yamaguchi University Graduate School of Medicine, 1-1-1 Minamikogushi, Ube, Yamaguchi, 755-8505, Japan; Kokoro no Hospital Machida, 2140 Kamioyamadamachi, Machida, Tokyo 194-0201, Japan

**Keywords:** Major depressive disorders, Biomarkers, Extracellular vesicles, *N*-linked glycosylation, von Willebrand factor

## Abstract

The clinical diagnosis of major depressive disorder (MDD), a heterogeneous disorder, still depends on subjective information in terms of various symptoms regarding mood. Detecting extracellular vesicles (EVs) in blood may result in finding a diagnostic biomarker that reflects the depressive stage of patients with MDD. Here, we report the results on the glycosylation pattern of enriched plasma EVs from patients with MDD and age-matched healthy subjects. In this cohort, the levels of *Triticum vulgaris* (wheat germ) agglutinin (WGA), *N*-acetyl glucosamine (GlcNAc) and *N*-acetylneuraminic acid (Neu5Ac, sialic acid) - binding lectin, were significantly decreased in patients with MDD in depressive state compared to healthy subjects (area under the curve (AUC): 0.87 (95% confidence interval (CI) 0.76 - 0.97)) and in remission state (AUC: 0.88 (95% CI 0.72 - 1.00)). Furthermore, proteome analysis revealed that the von Willebrand factor (vWF) was a significant factor recognized by WGA. WGA-binding vWF antigen differentiated patients with MDD versus healthy subjects (AUC: 0.92 (95% CI 0.82 - 1.00)) and the same patients with MDD in depressive versus remission state (AUC: 0.98 (95% CI 0.93 - 1.00)). In this study, the change patterns in the glycoproteins contained in plasma EVs support the usability of testing to identify patients who are at increased risk of depression during antidepressant treatment.

## Introduction

Depression is a major mental health challenge globally and the leading cause of mental health-related disability worldwide (Herrman et al., 2019). Even among international consensus diagnostic criteria, such as the International Classification of Diseases 11th (ICD-11) and the Diagnostic and Statistical Manual of Mental Disorders, Fifth Edition Text Revision (DSM-5-TR), diagnosing depression is based on a combination of symptoms. These diagnostic methods do not include objective laboratory findings (Malhi & Mann, 2018). Diagnosis and symptom evaluation of major depressive disorder (MDD) have depended on subjective information such as physicians’ clinical experiences and patient self-assessments, which can unfortunately lead to misdiagnosis (Mitchell et al., 2009). Therefore, it is highly desirable to elucidate the pathophysiology of depression and discover diagnostic markers for depression that reflect the pathophysiology of depression to change this situation.

Extracellular vesicles (EVs) are microscale vesicles composed of lipid bilayers released by various cell types, containing proteins, nucleic acids, lipids, and metabolites (Tominaga, 2021). Since EVs are involved in intercellular signaling, as they are transported and function between host and recipient cells (Xu et al., 2016), EVs are valuable tools for studying cell-cell communication in biological processes, including cancer, autoimmune disease, and neurodegenerative disorder, leading to blood-based biomarkers for early diagnosis disease (Wu et al., 2020; Delpech et al., 2019). For a diagnosis of MDD, most preclinical studies focus on proteins and microRNAs in EVs related to metabolic pathways in the central nervous system (CNS), neuro-inflammation, and neuroplasticity in the development of MDD (Li et al, 2021; Gelle et al., 2021; Hung et al., 2021). L1 cell adhesion molecule (L1CAM/CD171) is a helpful marker protein of EVs with putative CNS neuronal origin (Goetzl et al., 2021; Saeedi et al., 2021; Nasca et al., 2021). However, recent reports have questioned L1CAM as a marker protein from brain neuronal EVs because L1CAM is ubiquitously and transiently expressed in immune cells, specific epithelial cells, endothelial cells, and neural cells (Norman et al., 2021). Moreover, since there is still no standardized extraction protocol to purify EVs from peripheral blood for a diagnosis of MDD, the abundance of EVs and key factors in EVs can make a significant difference in each experiment and across facilities.

Glycosylated proteins, lipids, neurotransmitters, and hormones have important roles in communicating with surrounding cells through discrimination of their biomolecules (Ohtsubo & Marth, 2006). These glycosylated molecules are promising disease biomarkers (Kronimus et al., 2023; Pradeep et al., 2023; Paton et al., 2021; Chandler & Goldman, 2013; Costa et al., 2023). N-glycosylation plays a crucial role in several biological processes, especially those in the immune system. Since the onset and course of MDD are associated with alterations in the immune response, several reports have indicated that N-linked glycosylation is relevant to the diagnosis of the depressive state in women and the antidepressant treatment response in serum and plasma (Boeck et al., 2018; Park et al., 2018). We also previously reported that glycan patterns determined by lectin array in plasma obtained from depression model mice and patients with MDD and that alterations in glycosylate structures could be a diagnostic marker (Yamagata et al., 2018). EVs also have glycoproteins and glycolipids on their surface layer and inside, and this glycosylation pattern changes according to the condition depending on biological processes and diseases (Nishida-Aoki et al., 2020). However, the importance of EV glycosylation in understanding depressive status remains largely unknown.

Therefore, this study investigated whether the glycosylation pattern of plasma EVs between patients with MDD in depressive or remission state and healthy subjects was different. Furthermore, we revealed the glycoprotein with characteristic glycans for patients with MDD to construct a novel detection system reflected in depressive status.

## Materials and Methods

### Human subjects

We carried out this study following the latest version of the Declaration of Helsinki. The Institutional Review Board of Yamaguchi University Hospital approved this study (H25-085-13, H23-153-19, H2022-203), and all subjects provided written informed consent for participation. We recruited patients with MDD and healthy subjects as previously described (Yamagata et al., 2018). Briefly, patients with MDD were recruited from Yamaguchi University Hospital or referred by clinics and hospitals in the area. All subjects were recruited between April 2012 and June 2013 and followed until August 2014. We screened patients with MDD and diagnosed them using a structured clinical interview that included the International Neuropsychiatric Interview [M.I.N.I.], Japanese version 5.0.0 (Otsubo et al., 2005). We defined depression by a score greater than 18 on the Hamilton Rating for Depression Scale (HDRS) (HAMILTON, 1960). The Global Assessment of Functioning Scale (GAF) was used to assess social functioning (Moos et al., 2000). The Mini-Mental State Examination (MMSE) was used to exclude patients with possible dementia (score < 24) (Folstein et al., 1975). We classified patients in remission following the DSM-5 criteria for full remission. Healthy subjects were recruited using advertisements in the local community and screened using the MINI and a clinical interview. Any healthy subjects with a family history of psychiatric disease were excluded from the study. The demographic data of patients with MDD and healthy subjects included in this study are summarized in Table S1.

### Isolation of EVs from human plasma

We collected venous fasting blood from subjects between 9:00 and 12:00. Blood samples were centrifuged at 2400 ×g for 5 min to separate plasma from peripheral blood cells. These samples were stored at −80 °C until we used them. 500 µL of processed plasma was directly overlaid onto qEV size exclusion columns (qEVoriginal-70 Gen2, Izon Science Ltd.), followed by sample concentration to a final volume of 400 µL. To estimate the protein concentration of each qEV fraction, we used the Micro BCA protein assay kit (Thermo Fisher Scientific Inc. MA, USA) according to the manufacturer’s protocol. To confirm the fractions of concentrated EVs, we did a western blot with antibodies against CD9 and CD63 as EV markers.

### Nanoparticle tracking analysis (NTA)

We measured the size distribution and concentration of particles with Videodrop (Myriade Paris, France). 7 µL of each EV fraction was used to measure the concentration and size of particles in each condition. The threshold was set at 4.2 for the removal of macro particles, and the exposure time was 0.90 ms for each frame. Accumulations were performed to count enough particles, with 100 - 300 particles counted for each condition.

### Lectin blotting

EVs were denatured with lysis buffer with 2-mercaptoethanol at 99°C for 5 min. Lysates were loaded 400 ng protein per lane to run SDS-PAGE. Separated proteins were transferred to the PVDF membrane and incubated with biotinylated lectins (#BK-1000, VECTOR LABORATORIES, INC.), following Table S2. After washing, streptavidin-HRP at 1:5000 dilution was incubated at room temperature for 20 min. The membrane was developed by Immobilon Western Chemiluminescent HRP Substrate and examined using an Amasham imaging system.

### Western blotting

EVs were denatured with SDS sample buffer with 100 mM of dithiothreitol (DTT) (Nacalai Tesque, Japan) at 99 °C for 5 min (for reduced) or without DTT at 37 °C for 30 min (for non-reduced). Lysates were loaded 400 ng protein per lane to run SDS-PAGE. We separated proteins by SDS-PAGE and transferred them to polyvinylidene fluoride (PVDF) membranes. The membranes were blocked with Block Ace (Bio-rad) and incubated overnight at 4°C with primary antibodies, which included anti-CD63 (3-13, 1:1000, Wako), anti-CD9 (12A12, 1:1000, Cosmo bio), anti-cytochrome C (1:1000, proteintech), anti-vWF (EPSISR15, 1:1000, abcam). After washing PBS containing 0.1% Tween 20, the membranes were incubated at room temperature for 1 hour with horseradish peroxidase (HRP)-conjugated anti-rabbit or anti-mouse IgG (CST) as secondary antibodies. Signals were developed using an Immobilon Western Chemiluminescent HRP Substrate (Cytiva) and examined using an Amasham imaging system (GE Healthcare). We used Fiji (ImageJ 1.53t) to analyze the intensity of bands.

### Deglycosylation enzyme treatment *in vitro* for plasma EVs

Plasma EVs (400 ng) of healthy subjects were treated with α2-3,6,8 Neuraminidase (Sialidase, P0720, New England BioLabs, MA, USA) at least 50 unit per sample at 37°C for 1 h. The removal of glycans under this condition was confirmed by lectin blot or ELISA. The particle tracking analysis confirmed that most EVs maintained their vesicle structures after the enzymatic digestion.

### Enzyme-linked immuno-sorbent assay (ELISA)

We equally mixed plasma EVs and M-PER (Thermo Scientific, MA, USA) to isolate proteins from plasma EVs for ELISA. We assessed samples with VWF Human ELISA Kit (#EHVWF, Invitrogen) according to the manufacturers’ instructions.

For constricting of the sandwich ELISA to quantify WGA-binding vWF, 96-well plates with the antibody against human vWF were treated with PNGase F (P0704, New England Biolabs, MA, USA) to remove N-linked glycan for 4 hours at 37 °C. After washing, plasma EVs were added to plates and incubated overnight at 4 °C. Plates were incubated with WGA lectin (1:1000) at room temperature for 1 hour, following diluted streptavidin-HRP room temperature for 45 min. We measured absorbance at 450 nm using a Flexstaion 3 (Molecular Devices, USA) within 30 min after adding the TMB Substrate.

### Mass spectrometry and proteomic analysis

We performed proteomic analysis as previously reported. The EV lysates were dissolved by an equal amount of M-PER buffer and suspended in SDS-PAGE sample buffer with dithiothreitol (DTT). The samples were boiled at 99 °C for 5 min and resolved by SDS-PAGE. The gel was stained using a Silver Stain KANTO III (Kanto Chemicals). After slicing the gel, sliced gels were incubated at 37°C for trypsin digestion. Recovered peptides were desalted by Ziptip c18 (Millipore). Samples were analyzed using a nanoLC/MS/MS system (DiNa HPLC system KYA TECH Corporation/QSTAR XL Applied Biosystems). Mass data acquisitions were piloted by Mascot search (MS/MS Ion Search).

### Transmission electron microscopy (TEM)

Formver/carbon coated copper grid (NISSIN EM co. ltd., Japan) was hydrophilized with JFC-1600 Auto Fine Coater (JEOL, Japan). 3μL of purified EVs in PBS were placed on the hydrophilized grid and absorbed for 3min. We washed the glid using 500 μL double-distilled H2O successive four drops, then negatively stained using 30 μL of 2.0% uranyl acetate successive four drops. The grid was air-dried after absorbing 2.0% uranyl acetate on the grid with filter paper. We imaged the grid with Tecnai G2 Spirit BioTWIN electron microscopy (FEI) operating 120 kV equipped with a Phurona CMOS camera (Emsis). After exporting the raw data images to TIFF format, we used Fiji (ImageJ 1.53t) for data analysis.

### Quantification and statistical analysis

We performed the statistical analysis and visualized data using R software (v4.3.1). We used Student’s t-test or Mann - Whitney U test to compare the differences between the two groups. We also used Bartlett’s test and′ one-way ANOVA for differences between the variances of the three groups. Graphs show mean ± standard division (SD). We used the pROC package (v1.18.5) in R software to estimate the prediction value for diagnosis. The area under the curve (AUC) of ROC was analyzed to judge the accuracy of the predictive model. We used the curve closest to the (0,1) to estimate the cut-off point.

## Results

### Characterization of plasma EVs from patients with major depressive disorders and healthy subjects

Participant demographic and clinical data acquired from the experiment are shown in Table S1. Differences in age, gender, HDRS, and GAF were nonsignificant among all groups. Differences in onset age and dose of antidepressants were also nonsignificant between depressive and remission states. We collected plasma EVs using qEV columns following a specific protocol (Fig. 1). EVs were purified from the plasma of healthy subjects (HS, n = 20) or patients with major depressive disorder (MDD, n = 21) through the size-exclusion chromatography (SEC). To collect the EVs enriched fraction, we performed a BCA protein assay to estimate protein concentration and western blotting with antibodies against CD63 and CD9 as EV-markers. Plasma proteins eluted later fractions (F10-12) than EVs (F7-9). Since both CD63 and CD9, specific EV markers, were detected using western blotting, we used fractions 7–9 in this experiment.

**Fig. 1.**
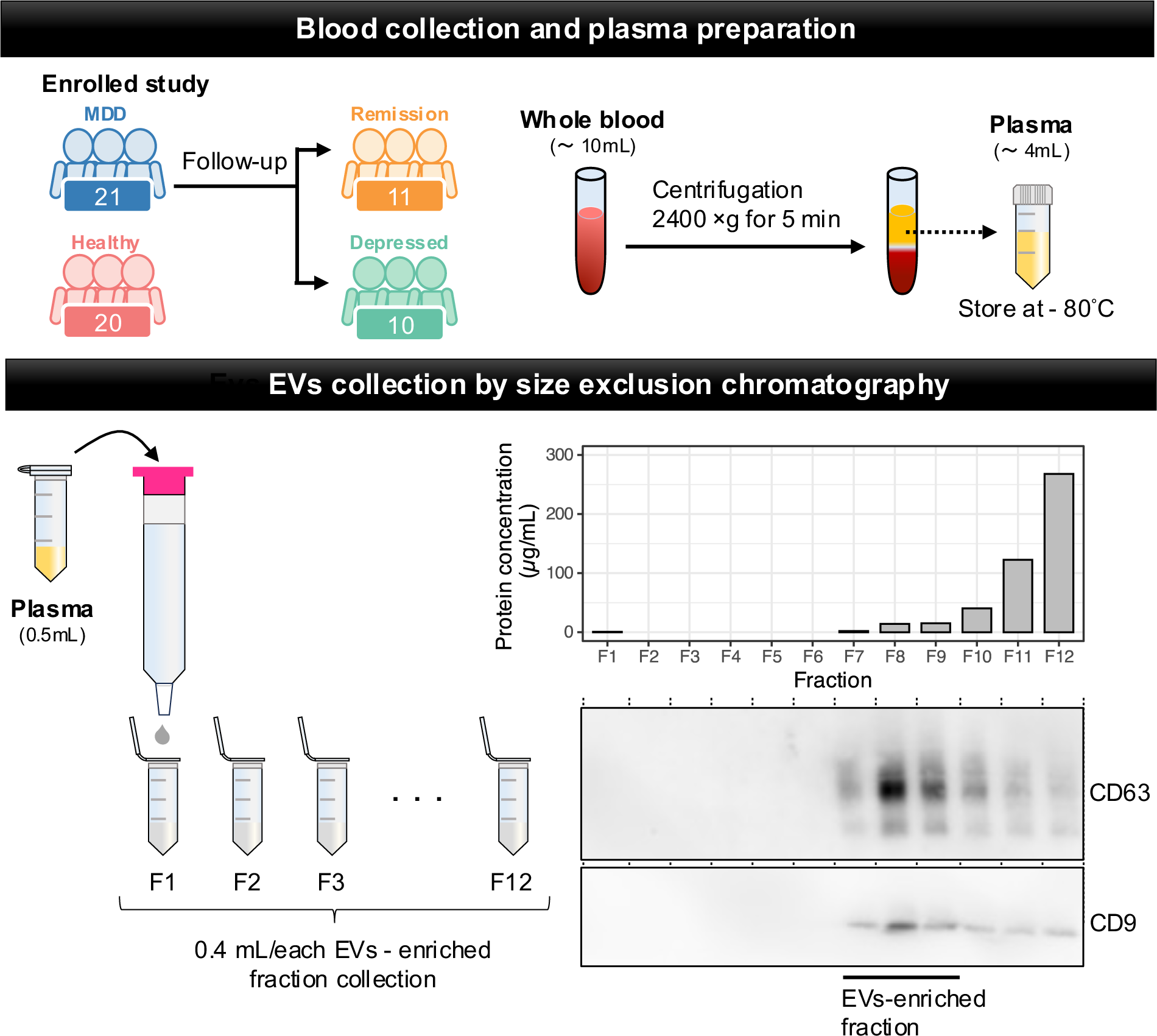
Overview of experimental framework for EVs enrichment and extraction by size-exclusion chromatography. We purified EVs from the plasma of healthy subjects (HS, n = 20) or patients with major depressive disorder (MDD, n = 21) by centrifugations, filtration, and passing through size-exclusion chromatography (SEC). The BCA protein assay and western blotting show a typical elution profile of plasma EVs obtained from a healthy subject.

To evaluate the characterizations of plasma EVs in depressive states, we isolated the fraction containing EVs obtained from plasma from both patients with MDD and healthy subjects by detecting the expression of the EV marker proteins, CD63 and CD9 (Fig. 2A). The purified EVs were observed by transmission electron microscopy (TEM), which showed the morphology, as commonly seen between patients with MDD and healthy subjects (Fig. 2B). Particle size distribution confirmed that the EVs from each sample population were in the range of 50 – 300 nm with peaks at around 150 nm (Fig. 2C). The modal particle size and the number of EV particles were similar among the EVs from healthy subjects and those in patients with MDD (Fig. 2D). On the other hand, the amount of protein in plasma EVs from patients with MDD was significantly decreased in that of healthy subjects. When we compared the modal particle size, the number of EV particles, and protein concentration between in depressive and remission states of the same patients, there was no statistically significant (Fig. 2E). These results indicated that protein contents in plasma EVs are possible to decrease or change during the progression of depression.

**Fig. 2.**
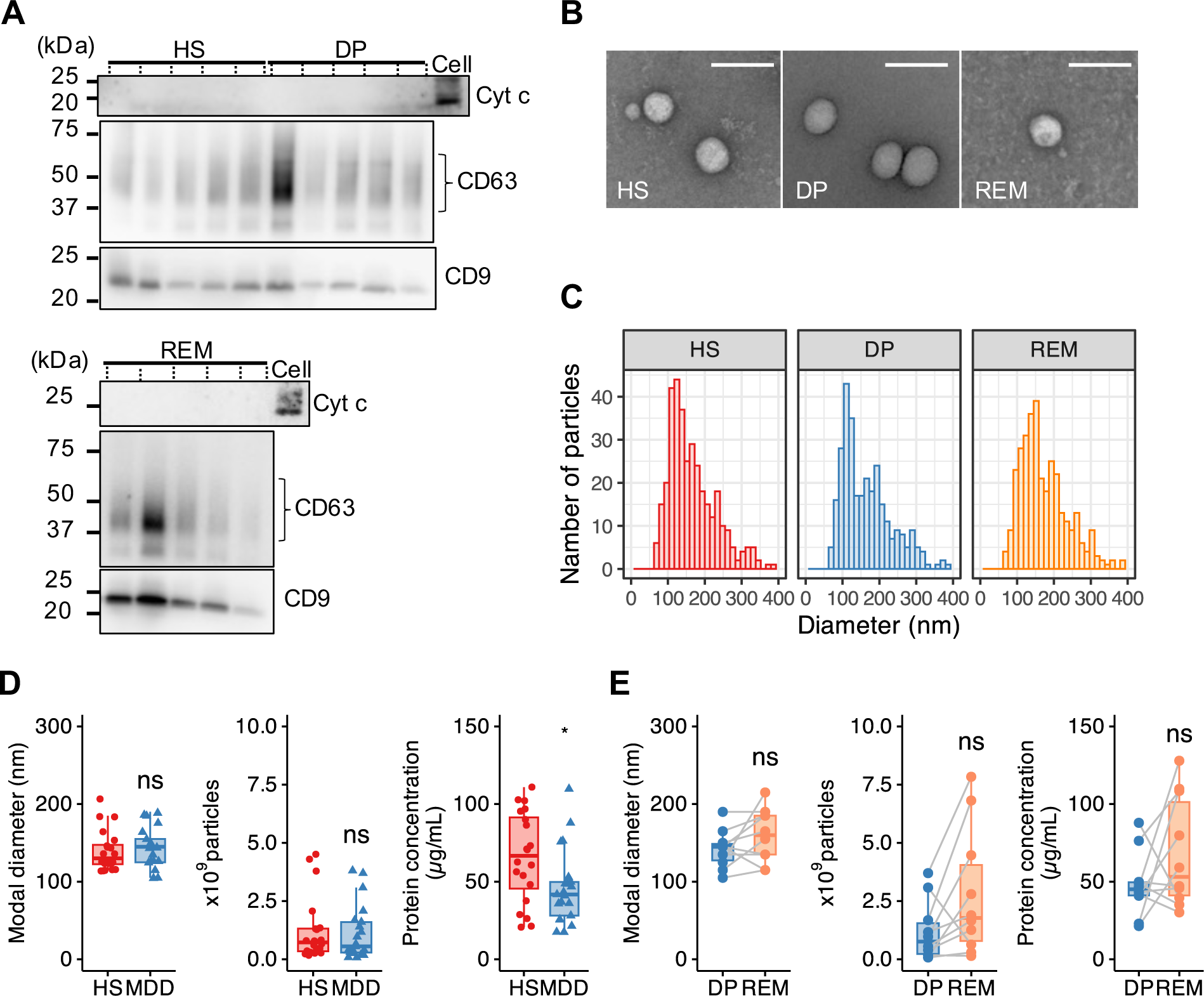
Efficient separation of EVs in plasma obtained from HS, DP, and REM by size exclusion chromatography. **(A)** Representative western blotting using plasma EVs in HS, patients with MDD in depressive state (DP), and remission state (REM) to confirm EVs-enriched fractions. We assessed protein extracts (400 ng) from cells or EVs using antibodies against EV protein markers (CD63, CD9) and a mitochondrial protein marker (cytochrome c (Cyt c)). We used protein extracts of HEK293T cells (Cell) to distinguish EVs from cells. **(B)** Representative photomicrographs of EVs isolated from plasma in HS, DP, or REM imaged by CryoEM. Scale bar: 100 nm. **(C)** Representative particle size histogram of EVs in the enriched plasma fractions in HS, DP, or REM by NTA. **(D)** Box plot of size, particle concentration, and protein concentration in plasma EVs after SEC. Each dot presents one sample. HS; n = 20, MDD: n = 21. **(E)** Paired box plots depicting individual patient data between patients with MDD (n = 10) in DP and REM in size, particle concentration, and protein concentration. We used Mann - Whitney U test or Student t-test to test for significance. * *p* < 0.05, n.s. nonsignificant.

### The extent of WGA-binding glycoprotein is reflected in the patients with MDD in a depressive state

To characterize EV glycosylation in patients with MDD in depressive state, we recognized glycans by lectin blotting. We summarized the glycan structures specific to each lectin in Table S2. SDS‒ PAGE revealed a complex mixture of proteins detected in plasma EVs when we loaded the same amount of EV protein. O-type glycosylation patterns among *Ulex europaeus* agglutinin 1 (UEA-1), *Arachis hypogaea* agglutinin (PNA), and *Dolichos biflorus* agglutinin (DBA) were rarely detected in both healthy subjects and patients with MDD in depressive state (Fig. 3A). On the other hand, N-type glycosylation patterns were detected with lectins recognized by Concanavalin A (ConA), *Ricinus communis* agglutinin (RCA120), and *Triticum vulgaris* (wheat germ) agglutinin (WGA), but not Soybean Agglutinin (SBA) (Fig. 3B). Significant differences observing for ConA, RCA120 and WGA between the two groups, we selected three lectins to measure the intensities of the visible bands at approximately 250 kDa. The amount of glycosylated protein with a molecular weight above 250 kDa recognized by WGA significantly decreased in plasma EVs from patients with MDD compared with healthy subjects (Fig. 3C, densitometric intensity of HS, 6221.34 ± 1422.17 and DP, 2833.87 ± 1254.72; p = 0.00727). Moreover, to validate the discovery results, we compared all participants in patients with MDD (n = 21) and healthy subjects (n = 20). The intensity of WGA was significantly lower in the patient with MDD than in the healthy subjects (Fig. 3D, densitometric intensity of HS, 5162.67 ± 3360.85 and DP, 1338.97 ± 1898.61; p = 0.00008). When we constructed a receiver operating characteristic curve (ROC) curve of relative intensity of WGA between patients with MDD in depressive state and healthy subjects, ROC analysis yielded an AUC of 0.87 (95% CI 0.76 - 0.97) (Fig. 3E), thus indicating the predictive power of plasma EVs with glycans in patients with MDD.

**Fig. 3.**
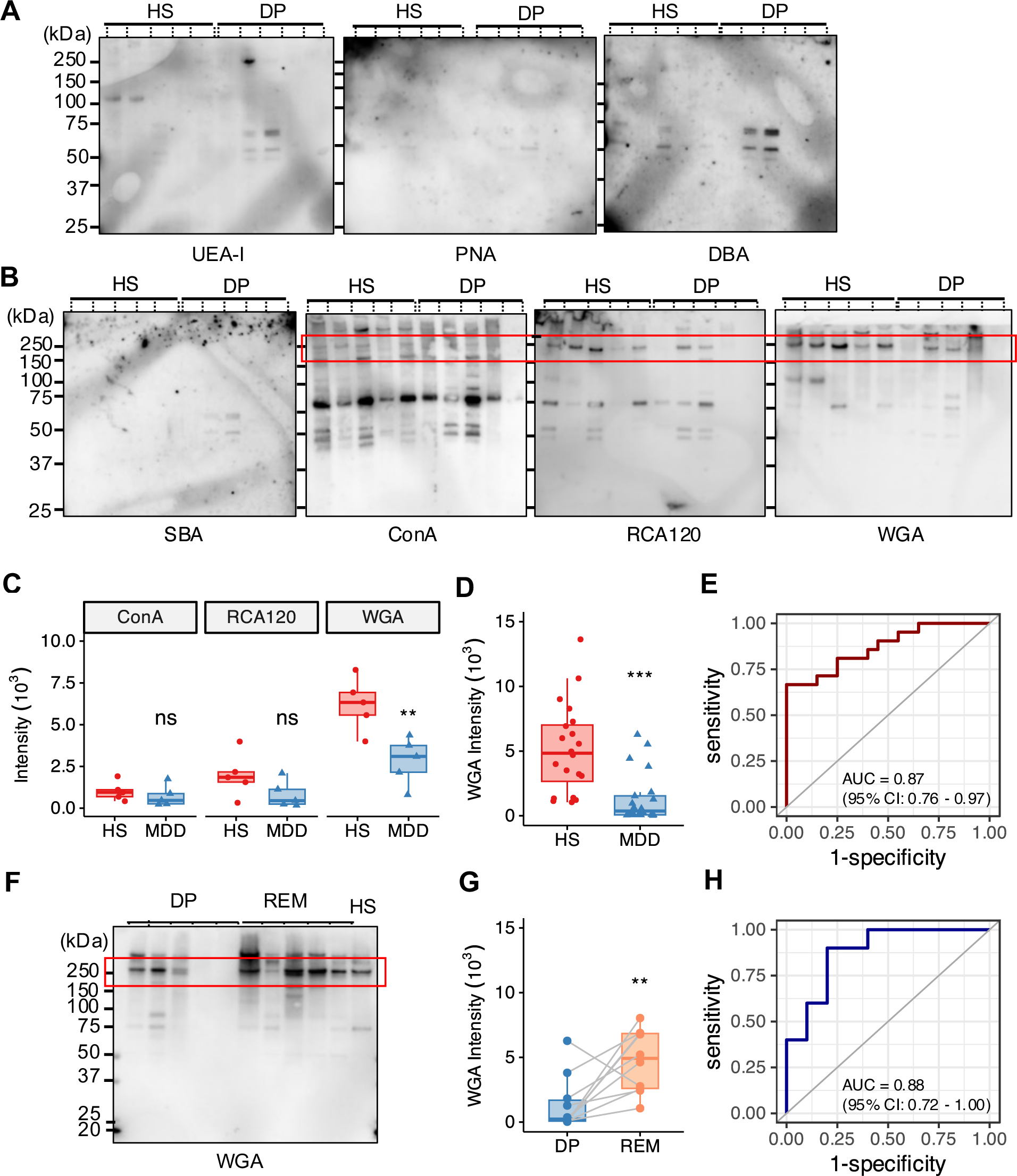
WGA lectin detects changes in the glycosylation status in plasma EVs in HS, MDD, and REM. **(A)** O-glycan profiling of plasma EVs of HS and DP by lectin blotting. We detected glycosyl chains by biotinylated lectins followed by HRP-conjugated streptavidin. Lectins used for detections: UEA-I, PNA, and DBA. We loaded same amount of protein concertation (400 ng/lane). **(B)** N-glycomic profiling of plasma EVs of HS and DP by lectin blotting. Lectins used for detections: SBA, ConA, RCA120, and WGA. We loaded same amount of protein concertation (400 ng/lane). The red line indicates bands at approximately 250 kDa. **(C)** we analyzed relative intensity in arbitrary units by Image J in (B). The number of independent: HS; n = 5, DP; n = 5. **(D)** The boxplots represent the signal intensity of protein bands in arbitrary units. **(E)** Receiver operating characteristic (ROC) curve analysis using the intensity of WGA lectin in plasma EVs to diagnose HS or MDD. A cutoff value is 2450 by the closest-top-left method. **(F)** Lectin blotting profiles detected by WGA in plasma EVs from patients with MDD in DP and REM. We used plasma EVs by HS as a positive control. The red line indicates bands at approximately 250 kDa. **(G)** Paired box plots depicting individual patient data between patients with MDD (n = 10) in DP and REM. **(H)** ROC curve analysis of intensity of WGA as a biomarker of DP compared with REM. A cutoff value is 2130 by the closest-top-left method. We used Mann - Whitney U test or Student t-test to test for significance. ** *p* < 0.01, *** *p* < 0.001, n.s. nonsignificant.

Next, to assess the diagnosis of depressive state, we compared the amount of WGA-binding protein with a molecular weight above 250 kD in plasma EVs between depressive and remission state of the same patients with MDD (n = 10) by lectin blotting (Fig. 3F). The intensity of the protein band at a molecular weight above 250 kDa in plasma EVs from depressive state individuals detected by WGA was significantly lower than that from EVs from those that had recovered to remission state (Fig. 3G, densitometric intensity of DP, 1390.40 ± 1993.23 and REM, 4723.40 ± 2277.96; p = 0.0040). In addition, the ROC curve showed an AUC of 0.88 (95% CI 0.72 - 1.00) for plasma EVs, which distinguished each status (Fig. 3H). We previously reported the results of a lectin array using leukocytes obtained from the human peripheral blood of patients with MDD and healthy subjects(Yamagata et al., 2018). Among them, there were no differences between the patients with MDD and healthy subjects determined by different sialic acid binding lectins, including ConA, RCA120, and WGA (Fig. S1). These data show that a decrease in glycoproteins recognized by WGA in plasma EV could indicate the status of depressive symptoms, which have the potential for diagnosis of depression.

### Proteome analysis reveals that vWF selectively contained in plasma EVs

The lectin of WGA can bind to oligosaccharides containing terminal N-acetyl glucosamine (GlcNAc) and N-acetylneuraminic acid (Neu5Ac, also known as sialic acid). To search for proteins that could link N-linked carbohydrate chains containing terminal GlcNAc and sialic acid, we first performed silver staining on samples from healthy subjects (Fig. 4A). We next analyzed the identified proteins by mass spectrometry. Among them, one of the most significant hits was the von Willebrand factor (vWF) (Fig. 4B, Fig. S2). vWF is a glycoprotein implicated in the maintenance of hemostasis at the sites of vascular injury by forming a molecular bridge between the subendothelial matrix and platelet-surface receptor complex (Peyvandi et al., 2011). We confirmed that the vWF proteins, detected by western blotting, had a molecular weight above 250 kDa in plasma EVs from healthy subjects and patients with MDD (Fig. 4C). Human vWF is capped by terminal negatively charged sialic acid residues (Ward et al., 2019). After sialidase digestion *in vitro*, the levels of the vWF protein decreased and shifted to a molecular weight of less than 250 kDa (Fig. 4D). Thus, our data suggested that plasma EVs contained vWF expressing terminal sialic acid.

**Fig. 4.**
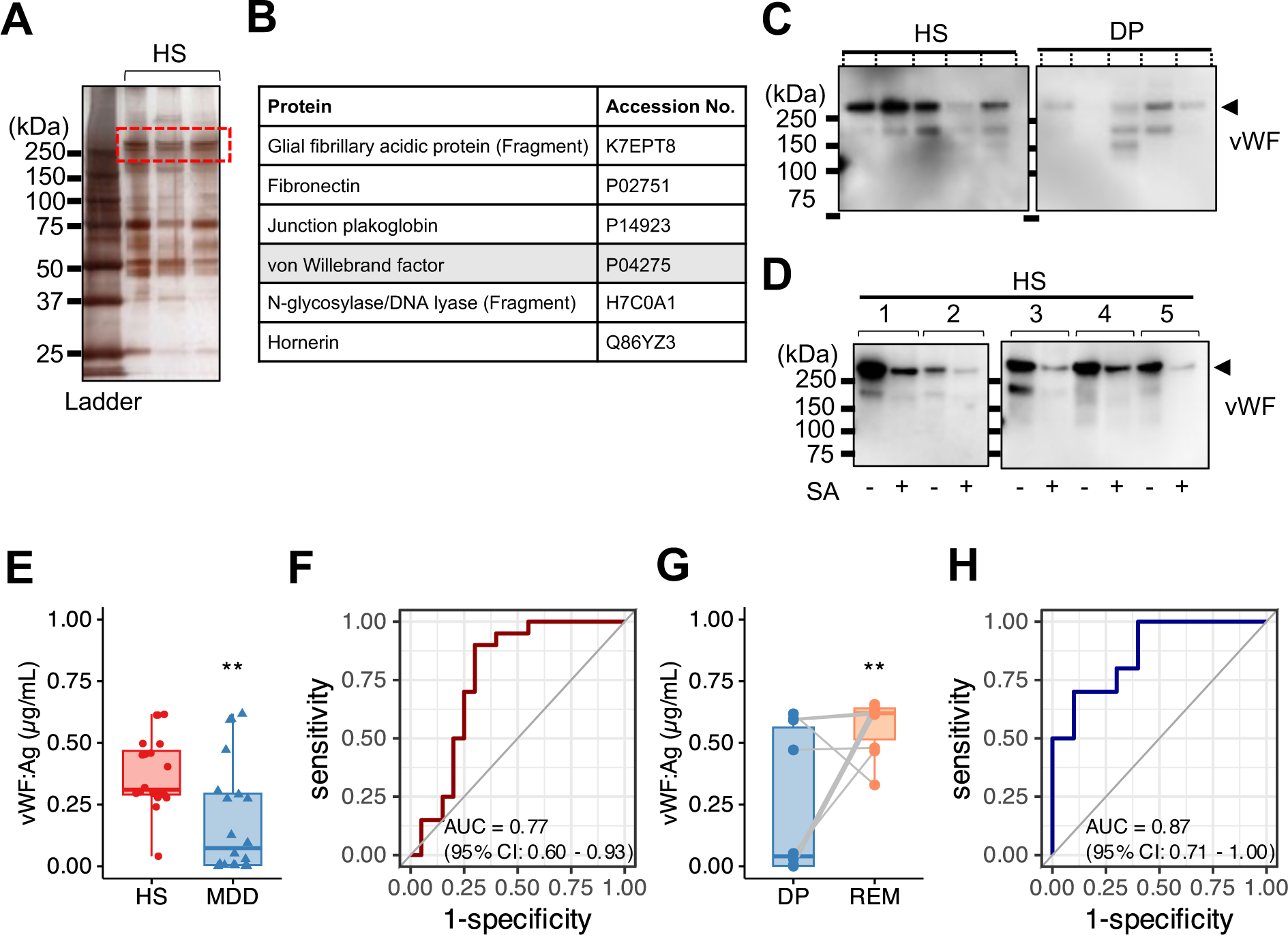
Proteome analysis revealed that vWF in plasma EVs related to the stage of MDD. **(A)** SDS-PAGE gel with silver staining. The gels contain protein standard (lane 1) and HS (lane 2 - 4). Coverage of above 250kDa protein (red dashed line) in plasma EV obtained HS analyzed by LC-MS. **(B)** List of polypeptides within above 250kDa protein identified in the proteomics analysis. **(C)** Representative image of western blotting for vWF protein (Molecular weight, 260 kDa) in plasma EVs (400 ng) obtained from HS (n = 5), patients with MDD in DP (n = 5). **(D)** Representative image of western blotting for plasma EVs proteins (400 ng) in HS (n = 5) with or without sialidase (SA) treatment. We detected the PVDF membrane strips with an antibody against vWF. **(E)** Densitometric measurement of vWF antigen (Ag) in plasma EVs from HS (n = 20) and MDD (n = 21) by sandwich ELISA. The absorbance read at 450 nm. **(F)** ROC curve analysis using vWF Ag in plasma EVs for diagnosis of MDD. A cutoff value is 0.276 µg/mL by the closest-top-left method. **(G)** Paired box plots depicting individual patient data between patients with MDD (n = 10) in DP and REM. **(H)** ROC curve analysis using vWF Ag in plasma EVs for diagnosis of MDD. A cutoff value is 0.606 µg/mL by the closest-top-left method. We used Mann - Whitney U test or Student t-test to test for significance. ** *p* < 0.01.

To assess the expression of vWF protein in patients with MDD and healthy subjects, we performed sandwich ELISA on the vWF protein from the same amount of plasma EVs in each sample. vWF protein expression was significantly lower in plasma EVs from patients with MDD in depressive state (mean concentration 0.187 ± 0.220 µg/mL, n = 21) than in those from healthy subjects (mean concentration 0.376 ± 0.143 µg/mL, n = 20) (Fig. 4E). ROC analysis of plasma EVs from patients with MDD in depressive state and healthy subjects yielded an AUC of 0.77 (95% CI 0.60 – 0.93) (Fig. 4F). Furthermore, the concentration of vWF protein in plasma EVs recovered during remission status (mean concentration 0.572 ± 0.104 µg/mL, n = 10) (Fig. 4G). In validation, vWF protein in depressive state was distinguished from remission status by ROC (AUC of 0.87 (95% CI 0.71 - 1.00)) (Fig. 4H). Hence, these results indicate that vWF-containing plasma EVs strongly correlated with the patients with MDD in depressive state.

Plasma consistently contains abundant vWF because multimer vWF is essential in immediately responding to an injury (Peyvandi et al., 2011). The amount of released vWF to plasma was no significant difference between patients with MDD (3.979 ± 0.683 µg/mL, n = 10) and healthy subjects (4.461 ± 0.480 µg/mL, n = 10) (Fig. S3). We could confirm that the amount of vWF on plasma EVs was altered in patients with MDD compared to healthy subjects, which did not depend on the amount of vWF in plasma.

### The expression levels of WGA-vWF strongly correlate with the treatment state in patients with MDD

To determine if plasma EVs containing WGA-binding vWF (WGA-vWF) could be applied as a diagnostic marker for MDD in the clinic, we designed a sandwich ELISA system to detect both WGA and vWF antigen in plasma EVs (Fig. 5A). The anti-vWF antibody without N-linked carbohydrate using PNGase F was immobilized on a 96-well plate. Plasma EVs were captured and detected with biotinylated WGA. The constructed standard curve exhibited concentration-dependent linearity (Fig. S4). When we also measured the absorbance of WGA-binding vWF after sialidase treatment, the absorbance was decreased compared with no treatment (Fig. 5B). Hence, the ELISA we developed was acceptable for detecting both WGA and vWF in plasma EVs.

**Fig. 5.**
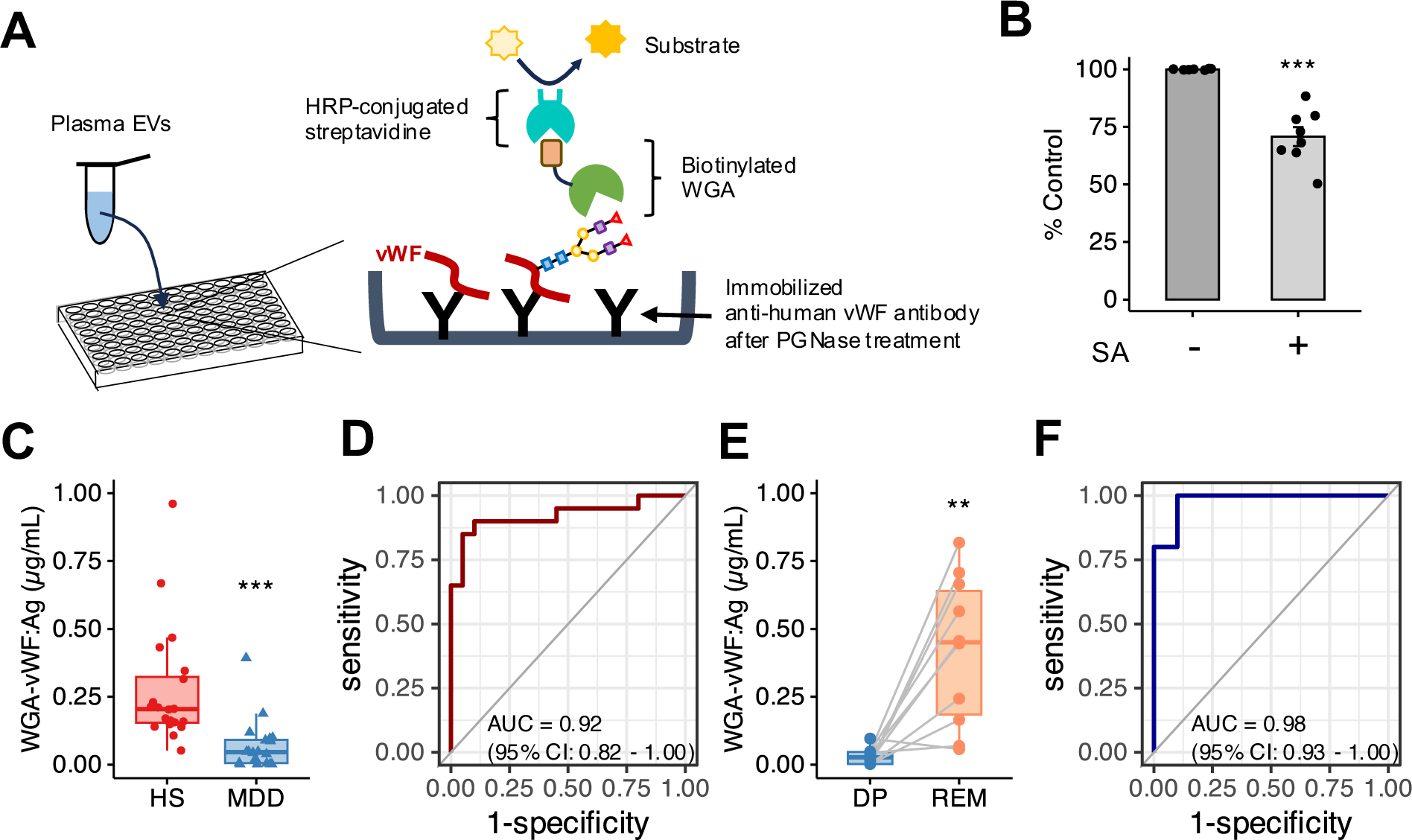
The levels of vWF Ag recognized by WGA in plasma EVs strongly correlate with the depressive state. **(A)** The shame of lectin-antibody sandwich ELISA for quantification of WGA-vWF protein. Bound vWF antibodies after PNGase treatment were detected using HRP-conjugated WGA lectin. The absorbance read at 450 nm. **(B)** Normalized absorbance of WGA-vWF antigen (Ag) in plasma EVs (1 µg protein each sample) in healthy subjects (n = 8) with or without SA treatment by the lectin-antibody sandwich ELISA. The level of vWF recognized by WGA showed significant changes due to the desialylation of vWF. **(C)** Quantification of WGA-vWF: Ag in plasma EVs by ELISA between patients with MDD (n = 21) and HS (n = 20). **(D)** ROC curve analysis of the concentration of WGA-vWF: Ag in plasma EVs for diagnosis of MDD in depressive state. The cutoff value is 0.129 µg/mL by the closest-top-left method. **(E)** Paired box plots depicting individual patient data of WGA-vWF: Ag in plasma EVs between patients with MDD (n = 10) in DP and REM. **(F)** ROC curve analysis of WGA-vWF: Ag in plasma EVs between DP and REM in the same patients with MDD. A cutoff value is 0.053 µg/mL by the closest-top-left method. We used Mann - Whitney U test or Student t-test to test for significance. ** *p* < 0.01, *** *p* < 0.001.

When we considered whether ELISA detected WGA-vWF in plasma EVs can help diagnose depression, WGA-vWF expression was significantly lower in plasma EVs of patients with MDD in depressive state (mean concentration 0.068 ± 0.104 µg/ml, n = 21) than those of healthy subjects (mean concentration 0.274 ± 0.212 µg/ml, n = 20) (Fig. 5C). ROC analysis indicated that the AUC value for the diagnosis was 0.92 (95% CI 0.82 - 1.00) between patients with MDD and healthy subjects (Fig. 5D). Furthermore, WGA-vWF expression remarkably increased from depressive (mean concentration 0.030 ± 0.029 µg/ml) to remission processes (mean concentration 0.419 ± 0.260 µg/ml) (Fig. 5E). We also could distinguish between patients with MDD in depressive and remission states (AUC of 0.98, 95% CI 0.93 - 1.00) (Fig. 5F). This ELISA combines WGA and vWF is more specific method than ELISA using the vWF antibody alone. Our findings suggested that WGA-vWF is a potential biomarker for the diagnosis of patients with MDD in depressive state.

## Discussion

In this study, we investigated the glycans on circulating EVs as potential signatures in patients with MDD and healthy subjects by lectin blotting. We found that the levels of *Triticum vulgaris* agglutinin (WGA), a lectin for terminal N-acetylglucosamine (GlcNAc) and N-acetylneuraminic acid (Neu5Ac, sialic acid), could distinguish not only between patients with MDD and healthy subjects but between patients with MDD in depressive state and remission. Additionally, we show that vWF containing sialic acid from circulating EVs in peripheral blood is a biomarker of patients with MDD. To our knowledge, this is the first report on a biomarker using EVs containing a glycoprotein that reflects the state of patients with MDD.

vWF is a large multidomain protein consisting of type A and D domains that can interact with multiple proteins (Gogia & Neelamegham, 2015; Denorme et al., 2019). vWF is mainly produced in endothelial cells and megakaryocytes, which are present in blood plasma (Leebeek & Eikenboom, 2016). vWF plays an important role in blocking blood vessels from binding to platelets via platelet receptors and aggregates after vascular injury (Bockenstedt et al., 1986; Ju et al., 2015; Bonazza et al., 2022). Multimer vWF builds in the Golgi complex and is stored within the Weibel-Palade body (WPB), similar to storage granules after being transported within lipid bilayers (Valentijn et al., 2008). vWF colocalizes with CD63, one of the specific EV markers, in the WPB of endothelial cells and is often associated with internal vesicles (Streetley et al., 2019). Accordingly, vWF may exist on EV surfaces, except for an adhesive surface or WPB on activated endothelial cells. Another group reported that vWF in plasma EVs obtained from patients with glioblastoma is a biomarker that might assist in diagnosing and managing glioblastoma (Sabbagh et al., 2021).

It is essential to determine whether vWF expression is the cause or origin of MDD symptoms in patients with MDD. Chronic stress leads to a tightly controlled process that involves a wide array of neuronal and endocrine systems(Smith & Vale, 2006). Depressed patients have hyperactivity of the hypothalamic-pituitary-adrenal axis (HPA) and high levels of glucocorticoids (GCs), which are secreted from the adrenal glands. The feedback to the brain, damages the portions of the brain environment, such as the hippocampus, resulting in a depressed mood (Ignácio et al., 2019). GCs bind to the promoter region of vWF and promote vWF expression, indicating a positive correlation between GC and vWF expression in Cushing syndrome patients (Casonato et al., 2008). Therefore, since vWF expression in plasma EVs significantly decreased in patients with MDD in depressive state, vWF does not appear to be downstream of the HPA system. Inflammation can also alter the brain signaling system through the CNS (Steinmetz & Turrigiano, 2010). Inflammatory cytokines, such as tumor necrosis factor-α (TNF-α), interleukin-1 beta (IL1β), and interleukin-6 (IL6), are secreted by microglia, which control the immune system in the brain (Steinmetz & Turrigiano, 2010; Zhu et al., 2006; Recasens et al., 2021) and activate serotonin transporters (Baganz & Blakely, 2013). Depressed patients have high expression levels of stress-induced inflammatory cytokines(Uddin et al., 2011). Multimer vWF secretion is high during inflammation associated with thrombus formation after vascular injury(B. Boneu et al., 1975), but vWF expression in vascular endothelial cells is decreased by TNF-α treatment in vitro (Li et al., 2015). Therefore, vWF expression in vascular endothelial cells may be downregulated by a stress response through an inflammatory pathway, leading to EV secretion with low levels of vWF from vascular endothelial cells to blood.

vWF monomers contain complex N-glycan and O-glycan structures (Ward et al., 2019). Glycan determinants play critical roles in regulating multiple aspects of vWF biology. Importantly, the vWF synthesized in endothelial cells is fully sialylated, whereas platelet-derived vWF has been shown to have significantly reduced vWF N-linked sialylation, which regulates proteolysis by ADAMTS13 (Mcgrath et al., 2013). Loss of sialic acid from glycans in the extremity in vWF terminates with Gal or GalNAc residues. Gal residues trigger enhanced vWF clearance through several different pathways, including the asialoglycoprotein receptor (ASGPR; also known as Ashwell-Morell receptor) on hepatocytes or macrophage galactose lectin receptor (MGL) on macrophages (Aguila et al., 2019). Thus, stabilizing vWF in blood to cover terminal sialic acid residues is important, leading to blood vessel formation, homeostasis, and repair after injury.

Finally, since MDD is a clinically heterogeneous phenotype, they are occasionally misdiagnosed at the expense of other mental disorders, such as bipolar disorder and schizophrenia (Mitchell et al., 2009). A comprehensive differential diagnosis of MDD is necessary for the clinical follow-up to distinguish the disorder from other mental conditions. Interestingly, the expression level of vWF in human plasma significantly increased in patients with bipolar disorder and schizophrenia compared to healthy subjects, with no differences between the two diagnostic groups (Hope et al., 2009). Endothelial hyperactivation is inferred to be an essential mechanism for the pathophysiology of bipolar disorder and schizophrenia. We showed that the low levels of vWF in plasma EVs in patients with MDD in depressive state have the potential for different characteristics from other mental disorders, leading to a rigorous tool for the clinical diagnosis of MDD. The blood-derived biomarker we found will contribute to the comprehensive diagnosis of MDD in addition to the psychological tests and the diagnostic imaging.

In summary, this study extensively screened the glycosylation patterns and glycoproteins expressed in plasma EVs from patients with MDD, and we identified sialylated vWF. This finding reveals not only a novel biomarker of MDD status but also original insight into the pathogenesis of MDD caused by endothelial dysfunction, which may provide novel approaches for MDD treatment.

## Data Availability

All data in this article (and its Supplementary Information files) are available. The other data analyzed in the current study are not publicly available for ethical reasons.

## Supplementary Materials

### Supplementary Figures

Fig. S1. Lectin array data of human leukocytes in healthy subjects (HS), patients with MDD in depressive (DP) or remission state (REM).

Fig. S2. Identification of vWF as a protein recognized by WGA.

Fig. S3. No significant differences for vWF levels in human plasma were found between patients with MDD and healthy subjects.

Fig. S4. Standard curve for a sandwich WGA-vWF ELISA.

### Supplementary Tables

Table S1. Demographics and clinical characteristics of study participants.

Table S2. Lectins used for detection of glycosylation of EVs.

## Acknowledgements

We thank participants and their generously volunteered time. We thank T. Matsubara, F. Higuchi, K. Harada, M. Masaaki for support with clinical protocol, patient care, or collection and provision of patient samples. We thank the Yamaguchi University Center for Gene Research for assistance with nanoparticle tracking analysis.

## Author contributions

N.Y., K.T., N.T. and S.N. conceived and designed the study. N.Y., K.T., C.N. and A.K performed experiments. N.Y. and K.T. did the data analysis and interpretation. N.Y., Y.M. and S.N. provided the clinical data and selected samples in this study. A.K. and S.N. did blood sample collection. Y.M. did transmission electron microscopy analysis. H.Y. did initial assay development and provided the lectin array data of human plasma. N.T. provided the technic and the knowledge in EV analyzing. N.Y., K.T., N.T. and S.N. wrote the manuscript and prepared the figures. All authors reviewed and edited the paper.

## Data and materials availabilidy

All data in this published article (and its Supplementary Information files) are available. The other data analyzed in the current study are not publicly available for ethical reasons.

