## Supplementary Materials for "Glycosylation state of vWF in circulating extracellular vesicles serves as a novel biomarker for predicting depression"

**Supplementary Figures**

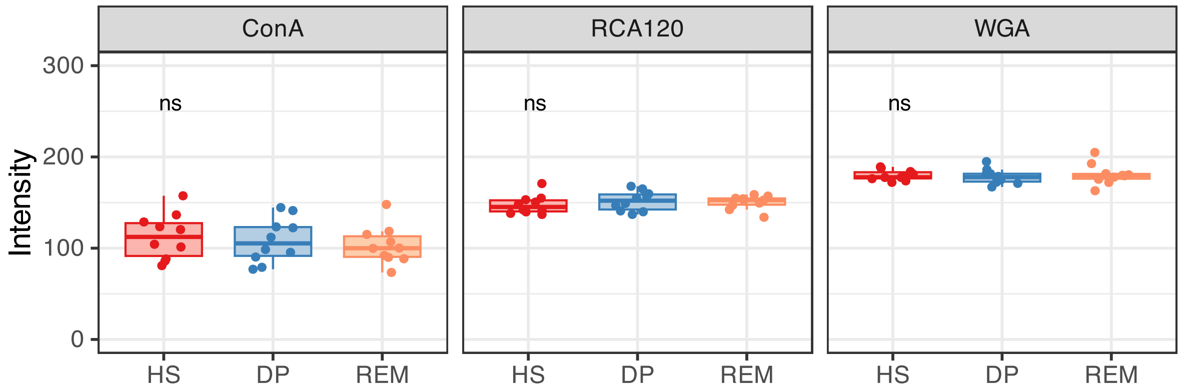


**Fig. S1. Lectin array data of human leukocytes in healthy subjects (HS), patients with MDD in depressive (DP) or remission state (REM).** Primary data are obtained from Yamagata H et al., 2017. Data are the mean ± s.d. Significance was determined by one-way ANOVA.

## A

| | | | HS #1 | | HS #2 | |
|---|---|---|---|---|---|---|
| Protein | Uniprot Accession | Molecula Weight (kDa) | Score | Matches | Score | Matches |
| Glial fibrillary acidic protein (Fragment) | K7EPT8 | 50 | 82 | 3(2) | 50 | 1(1) |
| Fibronectin | P02751 | 230 | - | - | 50 | 1(1) |
| Junction plakoglobin | P14923 | 23 | 37 | 2(1) | - | - |
| von Willebrand factor | P04275 | 260 | 36 | 3(1) | 53 | 5(1) |
| N-glycosylase/DNA lyase (Fragment) | H7C0A1 | | - | - | 34 | 3(0) |
| Hornerin | Q86YZ3 | 50 | 20 | 2(0) | - | - |

## B

```
>sp|P04275|VWF_HUMAN von Willebrand factor OS=Homo sapiens OX=9606 GN=VWF PE=1 SV=4
MIPARFAGVL LALALILPGT LCAEGTRGRS STARCSLFGS DFVNTFDGSM YSFAGYCSYL LAGGCQKRSF SIIGDFQNGK RVSLSVYLGE FFDIHLFVNG
TVTQGDQRVS MPYASKGLYL ETEAGYYKLS GEAYGFVARI DGSGNFQVLL SDRYFNKTCG LCGNFNIFAE DDFMTQEGTL TSDPYDFANS WALSSGEQWC
ERASPPSSSC NISSGEMQKG LWEQCQLLKS TSVFARCHPL VDPEPFVALC EKTLCECAGG LECACPALLE YARTCAQEGM VLYGWTDHSA CSPVCPAGME
YRQCVSPCAR TCQSLHINEM CQERCVDGCS CPEGQLLDEG LCVESTECPC VHSGKRYPPG TSLSRDCNTC ICRNSQWICS NEECPGECLV TGQSHFKSFD
NRYFTFSGIC QYLLARDCQD HSFSIVIETV QCADDRDAVC TRSVTVRLPG LHNSLVKLKH GAGVAMDGQD VQLPLLKGDL RIQHTVTASV RLSYGEDLQM
DWDGRGRLLV KLSPVYAGKT CGLCGNYNGN QGDDFLTPSG LAEPRVEDFG NAWKLHGDCQ DLQKQHSDPC ALNPRMTRFS EEACAVLTSP TFEACHRAVS
PLPYLRNCRY DVCSCSDGRE CLCGALASYA AACAGRGVRV AWREPGRCEL NCPKGQVYLQ CGTPCNLTCR SLSYPDEECN EACLEGCFCP PGLYMDERGD
CVPKAQCPCY YDGEIFQPED IFSDHHTMCY CEDGFMHCTM SGVPGSLLPD AVLSSPLSHR SKRSLSCRPP MVKLVCPADN LRAEGLECTK TCQNYDLECM
SMGCVSGCLC PPGMVRHENR CVALERCPCF HQGKEYAPGE TVKIGCNTCV CQDRKWNCTD HVCDATCSTI GMAHYLTFDG LKYLFPGECQ YVLVQDYCGS
NPGTFRILVG NKGCSHPSVK CKKRVTILVE GGEIELFDGE VNVKRPMKDE THFEVVESGR YIILLLGKAL SVVWDRHLSI SVVLKQTYQE KVCGLCGNFD
GIQNNDLTSS NLQVEEDPVD FGNSWKVSSQ CADTRKVPLD SSPATCHNNI MKQTMVDSSC RILTSDVFQD CNKLVDPEPY LDVCIYDTCS CESIGDCACF
CDTIAAYAHV CAQHGKVVTW RTATLCPQSC EERNLRENGY ECEWRYNSCA PACQVTCQHP EPLACPVQCV EGCHAHCPPG KILDELLQTC VDPEDCPVCE
VAGRRFASGK KVTLNPSDPE HCQICHCDVV NLTCEACQEP GGLVVPPTDA PVSPTTLYVE DISEPPLHDF YCSRLLDLVF LLDGSSRLSE AEFEVLKAFV
VDMMERLRIS QKWVRVAVVE YHDGSHAYIG LKDRKRPSEL RRIASQVKYA GSQVASTSEV LKYTLFQIFS KIDRPEASRI TLLLMASQEP QRMSRNFVRY
VQGLKKKKVI VIPVGIGPHA NLKQIRLIEK QAPENKAFVL SSVDELEQQR DEIVSYLCDL APEAPPPTLP PDMAQVTVGP GLLGVSTLGP KRNSMVLDVA
FVLEGSDKIG EADFNRSKEF MEEVIQRMDV GQDSIHVTVL QYSYMVTVEY PFSEAQSKGD ILQRVREIRY QGGNRTNTGL ALRYLSDHSF LVSQGDREQA
PNLVYMVTGN PASDEIKRLP GDIQVVPIGV GPNANVQELE RIGWPNAPIL IQDFETLPRE APDLVLQRCC SGEGLQIPTL SPAPDCSQPL DVILLLDGSS
SFPASYFDEM KSFAKAFISK ANIGPRLTQV SVLQYGSITT IDVPWNVVPE KAHLLSLVDV MQREGGPSQI GDALGFAVRY LTSEMHGARP GASKAVVILV
TDVSVDSVDA AADAARSNRV TVFPIGIGDR YDAAQLRILA GPAGDSNVVK LQRIEDLPTM VTLGNSFLHK LCSGFVRICM DEDGNEKRPG DVWTLPDQCH
TVTCQPDGQT LLKSHRVNCD RGLRPSCPNS QSPVKVEETC GCRWTCPCVC TGSSTRHIVT FDGQNFKLTG SCSYVLFQNK EQDLEVILHN GACSPGARQG
CMKSIEVKHS ALSVELHSDM EVTVNGRLVS VPYVGGNMEV NVYGAIMHEV RFNHLGHIFT FTPQNNEFQL QLSPKTFASK TYGLCGICDE NGANDFMLRD
GTVTTDWKTL VQEWTVQRPG QTCQPILEEQ CLVPDSSHCQ VLLLPLFAEC HKVLAPATFY AICQQDSCHQ EQVCEVIASY AHLCRTNGVC VDWRTPDFCA
MSCPPSLVYN HCEHGCPRHC DGNVSSCGDH PSEGCFCPPD KVMLEGSCVP EEACTQCIGE DGVQHQFLEA WVPDHQPCQI CTCLSGRKVN CTTQPCPTAK
APTCGLCEVA RLRQNADQCC PEYECVCDPV SCDLPPVPHC ERGLQPTLTN PGECRPNFTC ACRKEECKRV SPPSCPPHRL PTLRKTQCCD EYECACNCVN
STVSCPLGYL ASTATNDCGC TTTTCLPDKV CVHRSTIYPV GQFWEEGCDV CTCTDMEDAV MGLRVAQCSQ KPCEDSCRSG FTYVLHEGEC CGRCLPSACE
VVTGSPRGDS QSSWKSVGSQ WASPENPCLI NECVRVKEEV FIQQRNVSCP QLEVPVCPSG FQLSCKTSAC CPSCRCERME ACMLNGTVIG PGKTVMIDVC
TTCRCMVQVG VISGFKLECR KTTCNPCPLG YKEENNTGEC CGRCLPTACT IQLRGGQIMT LKRDETLQDG CDTHFCKVNE RGEYFWEKRV TGCPPFDEHK
CLAEGGKIMK IPGTCCDTCE EPECNDITAR LQYVKVGSCK SEVEVDIHYC QGKCASKAMY SIDINDVQDQ CSCCSPTRTE PMQVALHCTN GSVVYHEVLN
AMECKCSPRK CSK
```

**Fig. S2. Identification of vWF as a protein recognized by WGA. (A)** Mass spectrometry-based identification of von Willebrand factor (vWF) in plasma EVs obtained from healthy subjects. (**B)** Sequence of human vWF (Uniplot: P04275) with the peptides identified by mass spectrometry indicated in red.

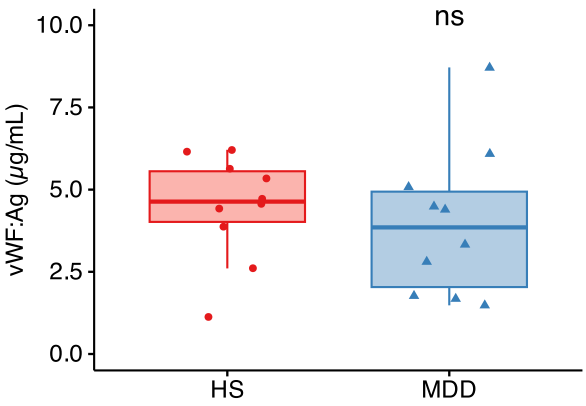


**Fig. S3. No significant differences for vWF levels in human plasma were found between patients with MDD and healthy subjects.** Quantification of vWF antigen (Ag) by sandwich ELISA between patients with MDD in depressive state (MDD, n = 10) and HS (n = 10). Significance was determined by Mann - Whitney U test to test for significance. n.s. nonsignificant.

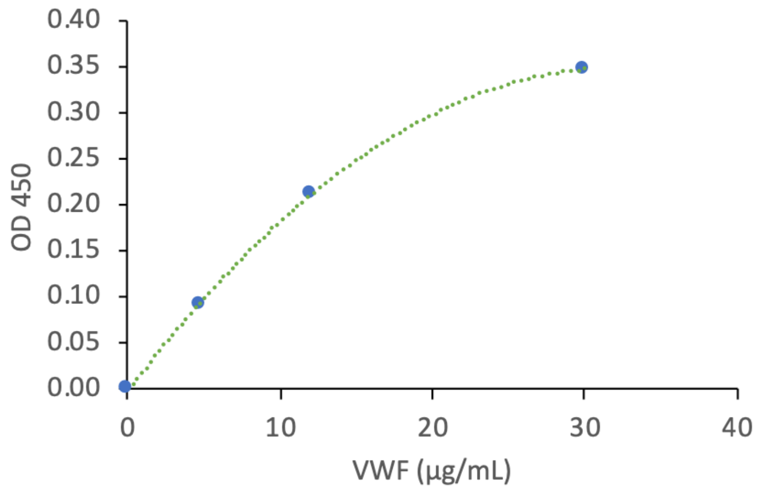


**Fig. S4. Standard curve for a sandwich WGA-vWF ELISA.** Absorbance (OD450) correlates with the amount of human recombinant vWF. Graph shows average values of technical duplicates on different days. R2 = 0.9998.

**Supplementary Tables**

**Table S1. Demographics and clinical characteristics of study participants.**

| | **Baseline** | | | **After follow-up in MDD cohort** | | |
|---|---|---|---|---|---|---|
| | HS | MDD | *p* - value | MDD-DP | MDD-REM | *p* - value |
| Sample size | 20 | 21 | | 10 | 11 | |
| Age (y) | 55.5 ± 7.6 | 57.6 ± 6.0 | 0.249* | 60.2 ± 5.5 | 56.5 ± 5.5 | 0.173* |
| Male/Female | 7/13 | 9/12 | 0.606† | 5/5 | 4/7 | 0.528† |
| Onset age (y) | - | 49.0 ± 12.7 | - | 53.0 ± 11.3 | 45.5 ± 13.4 | 0.217* |
| HDRS | 1.1 ± 1.0 | 23.8 ± 3.6 | $9.85e^{-27}$* | 25.0 ± 4.3 | 3.3 ± 2.6 | $1.43e^{-10}$* |
| GAF | 92.0 ± 4.3 | 51.2 ± 6.7 | $6.64e^{-24}$* | 49.5 ± 7.5 | 86.5 ± 7.8 | $6.52e^{-9}$* |
| Antidepressants§ (equivalent dose of imipramine) (mg) | - | 220.1 ± 104.8 | - | 190.0 ± 88.9 | 209.3 ± 116.6 | 0.691* |

Demographic data of healthy subjects (HS) and patients with major depressive disorder (MDD). After the follow-up period in the MDD cohort, we determined a diagnosis of permanent depressive (MDD-DP) or remission state (MDD-REM). Data is presented as mean (standard deviation) or the number of participants in each group. §point in time of diagnosis. *Independent-samples t-test. †Chi-squared test.

**Table S2. Lectins used for detection of glycosylation of EVs.**

| **Lectins** | **Origin** | **Primary recognition sugers** |
|---|---|---|
| **Con A** | Concanavalin A from *Canavalia ensiformis* | α-D-Man, α-D-Glc |
| **SBA** | *Soybean Agglutinin* Glycine max (soybean) | α<β GalNAc |
| **WGA** | *Triticum vulgaris (*wheat germ) agglutinin | D-GlcNAc, Neu5Ac |
| **DBA** | *Dolichos biflorus* agglutinin | α-GalNAc |
| **UEA-I** | *Ulex europaeus* agglutinin 1 | α-L-Fuc |
| **$RCA_{120}$** | *Ricinus communis* agglutinin | β-Gal |
| **PNA** | *Arachis hypogaea* (peanut) agglutinin | Gal-β (1-3)-GalNAc |

Fuc: L-Fucose, Gal: D-Galactose, GalNAc: *N*-Acetylgalactosamine, Glc: D-Glucose, GlcNAc: *N*-Acetylglucosamine, Neu5Ac: N-glycan *N*-acetylneuraminic acid
